# Research Letter: Association between long COVID symptoms and employment status

**DOI:** 10.1101/2022.11.17.22282452

**Authors:** Roy H. Perlis, Kristin Lunz Trujillo, Alauna Safarpour, Mauricio Santillana, Katherine Ognyanova, James Druckman, David Lazer

## Abstract

**Background:** Symptoms of Coronavirus-19 (COVID-19) infection persist beyond 2 months in a subset of individuals, a phenomenon referred to as long COVID, but little is known about its functional correlates and in particular the relevance of neurocognitive symptoms.

**Method:** We analyzed a previously-reported cohort derived from 8 waves of a nonprobability-sample internet survey called the COVID States Project, conducted every 4-8 weeks between February 2021 and July 2022. Primary analyses examined associations between long COVID and lack of full employment or unemployment, adjusted for age, sex, race and ethnicity, education, urbanicity, and region, using multiple logistic regression with interlocking survey weights.

**Results:** The cohort included 15,307 survey respondents ages 18-69 with test-confirmed COVID-19 at least 2 months prior, of whom 2,236 (14.6%) reported long COVID symptoms, including 1,027/2,236 (45.9%) reporting either ‘brain fog’ or impaired memory. Overall, 1,418/15,307 (9.3%) reported being unemployed, including 276/2,236 (12.3%) of those with long COVID and 1,142/13,071 (8.7%) of those without; 8,228 (53.8%) worked full-time, including 1,017 (45.5%) of those with long COVID and 7,211 (55.2%) without. In survey-weighted regression models, presence of long COVID was associated with being unemployed (crude OR 1.44, 95% CI 1.20-1.72; adjusted OR 1.23, 95% CI 1.02-1.48), and with lower likelihood of working full-time (crude OR 0.73, 95% CI 0.64-0.82; adjusted OR 0.79, 95% CI 0.70 -0.90). Among individuals with long COVID, the presence of cognitive symptoms – either brain fog or impaired memory – was associated with lower likelihood of working full time (crude OR 0.71, 95% CI 0.57-0.89, adjusted OR 0.77, 95% CI 0.61-0.97).

**Conclusion:** Long COVID was associated with a greater likelihood of unemployment and lesser likelihood of working full time in adjusted models. Presence of cognitive symptoms was associated with diminished likelihood of working full time. These results underscore the importance of developing strategies to respond to long COVID, and particularly the associated neurocognitive symptoms.

## INTRODUCTION

Symptoms of Coronavirus-19 (COVID-19) infection persist beyond 2 months in a subset of individuals, a phenomenon referred to as long COVID.^1^ This syndrome has become prevalent^2–4^, but little is known about its functional correlates and in particular the relevance of neurocognitive symptoms^5^. We therefore examined whether long COVID and neurocognitive symptoms in particular are associated with differential rates of employment as a proxy for functional impairment, to better guide development of potential interventions.

## METHOD

We analyzed a previously-reported cohort derived from 8 waves of a nonprobability-sample internet survey called the COVID States Project, conducted every 4-8 weeks between February 2021 and July 2022^2^. Respondents were consenting U.S. residents ages 18 and older, according to a protocol approved by the Institutional Review Board of Harvard University. We report results in accordance with AAPOR guidelines.

The cohort included all individuals who reported a positive COVID-19 test result, either PCR or antigen-based, at least 2 months prior to the survey month. Respondents were asked whether their acute symptoms had resolved; those who reported that they had not completed a checklist of commonly-reported symptoms. Respondents also self-reported sociodemographic variables including race and ethnicity based on U.S. Census categories, to allow for survey weighting.

Primary analyses examined associations between long COVID and lack of full employment or unemployment, adjusted for age, sex, race and ethnicity, education, urbanicity (urban, suburban, or rural), and region, using multiple logistic regression with interlocking survey weights in R 4.0.

## RESULTS

The cohort included 15,307 survey respondents ages 18-69 with test-confirmed COVID-19 at least 2 months prior, of whom 2,236 (14.6%) reported long COVID symptoms, including 1,027/2,236 (45.9%) reporting either ‘brain fog’ or impaired memory. Mean age was 38.8 (13.5) years; 9,678 (63.2%) identified as women and 5,629 (36.8%) men; 810 (5.3%) were Asian, 1,814 (11.9%) were Black, 1,501 (9.8%) were Hispanic, and 10,719 (70.0%) were White.

Overall, 1,418/15,307 (9.3%) reported being unemployed, including 276/2,236 (12.3%) of those with long COVID and 1,142/13,071 (8.7%) of those without; 8,228 (53.8%) worked full-time, including 1,017 (45.5%) of those with long COVID and 7,211 (55.2%) without. Of the remainder, 207 (14.8%) reported part-time or gig work, 13 (0.9%) being a homemaker, 53 (3.8%) being a student, and 11 (0.8%) being retired. In survey-weighted regression models, presence of long COVID was associated with being unemployed (crude OR 1.44, 95% CI 1.20-1.72; adjusted OR 1.23, 95% CI 1.02-1.48), and with lower likelihood of working full-time (crude OR 0.73, 95% CI 0.64-0.82; adjusted OR 0.79, 95% CI 0.70 -0.90). Among individuals with long COVID, the presence of cognitive symptoms – either brain fog or impaired memory – was associated with lower likelihood of working full time (crude OR 0.71, 95% CI 0.57-0.89, adjusted OR 0.77, 95% CI 0.61-0.97). (Figure 1).

**Figure 1.**
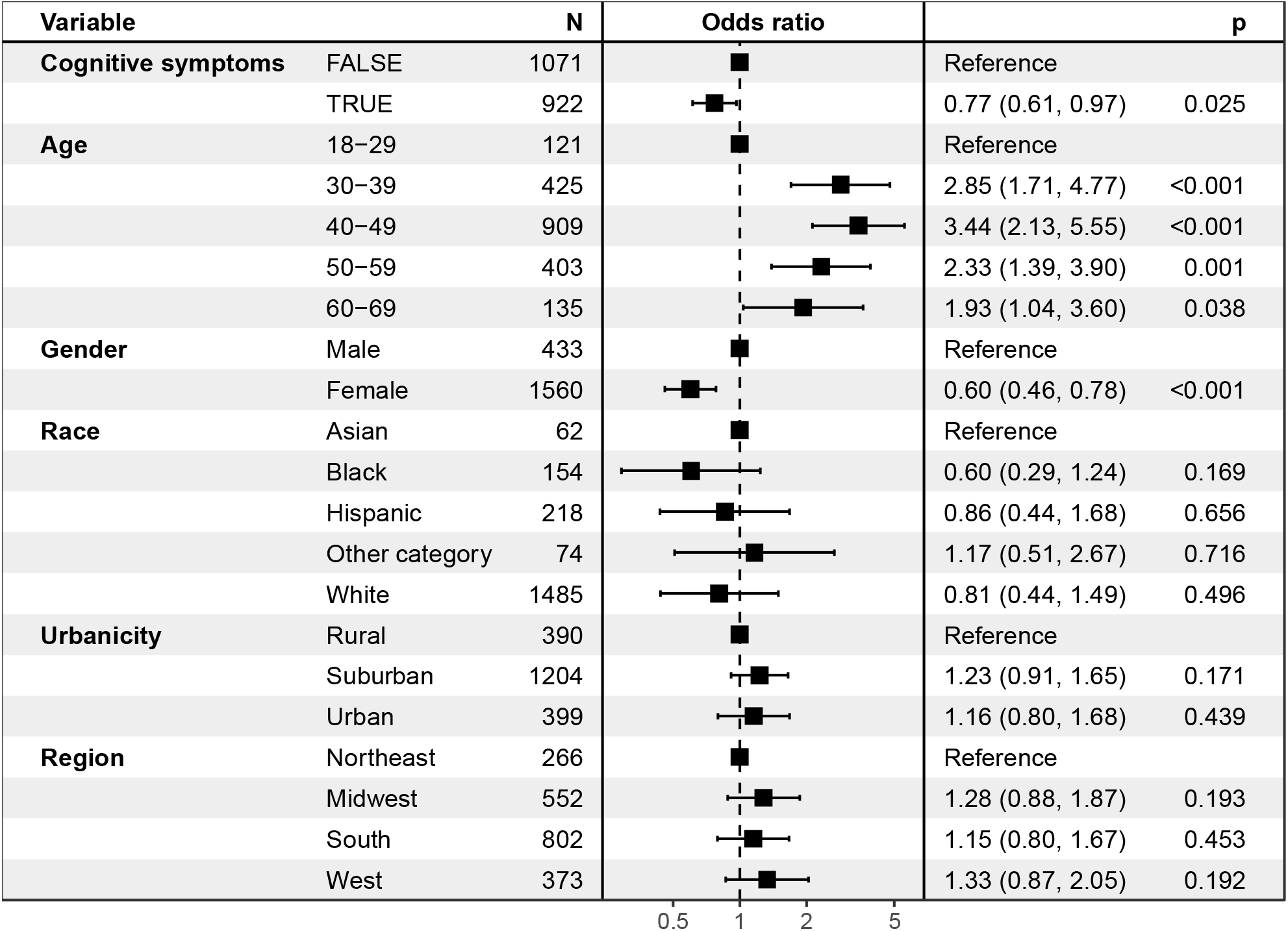
Among individuals with long COVID, symptoms associated with likelihood of not working full time at time of survey

## DISCUSSION

Among 15,307 US adults surveyed between February 2021 and March 2022, long COVID was associated with a greater likelihood of unemployment and lesser likelihood of working full time in adjusted models. Further, among those with long COVID, presence of cognitive symptoms was associated with diminished likelihood of working full time, extending recent reports associating cognitive symptoms with poor quality of life among employed individuals^5^.

A limitation in our analysis is the reliance on cross-sectional data, precluding determinations of causation. Moreover, the extent to which our results generalize remains to be determined, though prior work strongly suggests consistency with other approaches^6^.

These results underscore the importance of developing strategies to respond to long COVID, and particularly the associated neurocognitive symptoms. Whether rehabilitation strategies drawn from neurology and psychiatry can help to ameliorate the impact of such symptoms merits investigation. More generally, while the economic impact of the pandemic is difficult to estimate, these results suggest the importance of considering persistent effects of lost productivity.

## Data Availability

Data are not available

## Acknowledgements

The survey was supported in part by the National Science Foundation (Drs. Ognyanova, Lazer, Druckman) and by the National Institute of Mental Health (Drs. Perlis and Lazer -RF1MH132335). The sponsors did not have any role in design and conduct of the study; collection, management, analysis, and interpretation of the data; preparation, review, or approval of the manuscript; and decision to submit the manuscript for publication. The authors had the final responsibility for the decision to submit for publication. Dr. Perlis had full access to all the data in the study and takes responsibility for the integrity of the data and the accuracy of the data analysis.

## Author Contributions

Roy H. Perlis, MD MSc -- analyzed data, drafted and revised manuscript Kristin Lunz Trujillo, PhD – revised manuscript

Alauna Safarpour, PhD – revised manuscript

Mauricio Santillana, PhD – revised manuscript

Katherine Ognyanova, PhD – contributed to data collection and analysis, revised manuscript

James Druckman, PhD – revised manuscript

David Lazer, PhD – revised manuscript

